# Cohorting KPC+ *Klebsiella pneumoniae* (KPC-Kp) positive patients – a genomic exposé of cross-colonization hazards in a long-term acute care hospital (LTACH)

**DOI:** 10.1101/2020.02.07.20020669

**Authors:** Shawn E. Hawken, Mary K. Hayden, Karen Lolans, Rachel D. Yelin, Robert A. Weinstein, Michael Y. Lin, Evan S. Snitkin

## Abstract

**Objective:** Cohorting patients who are colonized or infected with multidrug-resistant organisms (MDROs) has been demonstrated to protect uncolonized patients from acquiring MDROs in healthcare settings. A neglected aspect of cohorting is the potential for cross-transmission within the cohort and the possibility of colonized patients acquiring secondary isolates with additional antibiotic resistance traits. We searched for evidence of cross-transmission of KPC+ *Klebsiella pneumoniae (*KPC-Kp) colonization among cohorted patients in a long-term acute care hospital (LTACH), and evaluated the impact of secondary acquisitions on resistance potential.

**Design:** Genomic epidemiological investigation

**Setting:** A high-prevalence LTACH during a bundled intervention that included cohorting KPC-Kp-positive patients.

**Methods:** Whole-genome sequencing (WGS) and location data were analyzed to identify potential cases of cross-transmission between cohorted patients.

**Results:** Secondary KPC-Kp isolates from 19 of 28 admission-positive patients were more closely related to another patient’s isolate than to their own admission isolate. In 14 of these 19 cases there was strong genomic evidence for cross-transmission (<10 SNVs) and the majority of these patients occupied shared cohort floors (12 cases) or rooms (5 cases) at the same time. Of the 14 patients with strong genomic evidence of acquisition, 12 acquired antibiotic resistance genes not found in their primary isolates.

**Conclusions:** Acquisition of secondary KPC-Kp isolates carrying distinct antibiotic resistance genes was detected in nearly half of cohorted patients. These results highlight the importance of healthcare provider adherence to infection prevention protocols within cohort locations, and motivate future studies to assess whether multiple-strain acquisition increases risk of adverse patient outcomes.

## Introduction

Cohorting of patients who are colonized or infected with high-priority healthcare pathogens has been demonstrated to prevent the spread of healthcare associated infections (HAIs).^1^ Cohorting works by physically separating colonized or infected patients together in one area for care, thereby preventing contact with other patients.^1^ In addition to being effective in outbreak settings,^2–5^ cohorting has been demonstrated to reduce cross-transmission in endemic healthcare settings with high colonization pressure, such as long-term acute care hospitals (LTACHs).^6,7^

Carbapenem resistant enterobacteriaceae (CRE) are multi-drug resistant organisms (MDROs) that are resistant to nearly all antibiotics and that are estimated to be responsible for 8,500 infections and 1,100 deaths in the U.S. annually.^8^ CRE have been labeled an urgent public health threat for nearly a decade, but despite wide-spread attention, infections with CRE have not decreased.^8^ Previous work has shown that LTACHs have a disproportionately high prevalence of CRE and that they likely contribute to transmission across regions.^9,10^ Encouragingly, a recent study demonstrated the effectiveness of a bundled intervention that included cohorting CRE-positive patients to reduce a particular type of CRE--*Klebsiella pneumoniae* that carry the KPC-type of carbapenemase (KPC-Kp)--in a LTACH with high KPC-Kp prevalence.^11^ This study highlights the potential for infection prevention interventions to reduce transmission in these complex and healthcare settings with a heavy burden of MDROs.^11^

Guidelines for preventing transmission in healthcare settings recommend placing “together in the same room (cohort) patients who are infected or colonized with the same pathogen” when single-patient rooms are unavailable.^1^ Yet molecular and phenotypic analyses of prominent healthcare pathogens like CRE indicate that strains of a given antibiotic resistance type are not necessarily equivalent in terms of resistance mechanisms and virulence genes.^12,13^ Cross-transmission of genetically diverse strains among cohorted patients could have clinically important consequences. First, patients are often treated empirically based on susceptibility results from prior cultures.^14–16^However, if a patient acquires new strains, this empiric antibiotic treatment strategy may fail because the secondary organism could carry different antibiotic resistance genes and therefore have a different susceptibility profile.^13,17,18^ Additionally, recent reports provide evidence in support of horizontal transfer of antibiotic resistance genes within patients,^19,20^ indicating that co-colonization with multiple strains can lead to entry of resistance genes into new genetic backgrounds.

Here, we examined the potential for multiple-strain colonization with KPC-Kp in a convenience sample of patients from a comprehensive surveillance study of KPC-Kp colonization in a Chicago LTACH.^11^ We hypothesized that by integrating whole-genome sequencing (WGS) and patient location data we would identify KPC-Kp colonized patients with evidence of acquisition of distinct secondary KPC-Kp strains through cross-transmission from other patients co-housed in cohort locations. Moreover, we predicted that secondary acquired strains would harbor antibiotic resistance genes that were not found in the patient’s admission isolate.

## Methods

### LTACH setting, study design and sample collection

Detailed information regarding the study design, intervention bundle and data collection are available in Hayden et. al 2015.^11^ Briefly and of relevance to the current manuscript, the study took place between 2011-2013 during a quality improvement project to prevent KPC-Kp colonization and infection in a Chicago LTACH where the average prevalence of KPC-Kp colonization was 30%. All location data and isolates presented here were collected from one LTACH during the intervention period,which included surveillance swab culture-based screening of all LTACH patients for KPC-Kp rectal colonization at LTACH admission and every two weeks (94% adherence), as well as efforts to separate KPC-Kp-positive and KPC-Kp-negative patients by placing KPC-Kp-positive patients in ward cohorts (91% adherence).^7^ Participating LTACHs deemed the study to be a quality improvement project and not research. The project was reviewed and determined to be a minimal-risk study by the institutional review board at Rush University Medical Center, which granted approval of the study along with a waiver of consent and Health Insurance Portability and Accountability Act waiver. ^11^

### Longitudinal convenience sample of KPC-Kp isolates from previously colonized patients

During the course of the original study, the first KPC-Kp surveillance isolate was collected from each colonized patient.^11^ Once a patient was found to be colonized with KPC-Kp, the patient was presumed to remain colonized indefinitely. Colonized patients were not rescreened systematically; however, additional ‘secondary’ KPC-Kp isolates were collected from a subset of patients whose prior colonization status was unclear to study staff at the time of screening.

The current analyses are restricted to this longitudinal, convenience sample of patients who were KPC-Kp positive at the study start or upon LTACH admission (within 3 days) and who also had one or more additional KPC-Kp surveillance isolates collected later. These ‘index’ patients were selected for study because they were housed in cohort locations during their entire LTACH stay, providing long periods of exposure to other KPC-Kp positive patients and potential opportunities for cross-transmission.

Among the index patients who had secondary isolates available, 100% were cohorted per-protocol: 21 patients with 46 secondary isolates shared a room with at least 1 patient who was KPC-Kp-positive before their secondary isolate being collected, and 8 patients with 15 secondary isolates did not have overlap with a positive patient before their secondary isolate was collected, but were instead housed in single patient rooms during the acquisition time frame for these isolates. Isolates from the 21 patients who shared a room with a putative KPC-Kp-positive donor prior to secondary acquisition were collected after patients shared a room with positive patients for a median of 51 days (range 1-132 days) prior to detection of a secondary isolate.

### Whole-genome sequencing

DNA was extracted with the MoBio PowerMag Microbial DNA kit and prepared for sequencing on an Illumina MiSeq instrument using the NEBNext Ultra kit and sample-specific barcoding. Library preparation and sequencing were performed at the Center for Microbial Systems at the University of Michigan or the University of Michigan Sequencing Core. Quality of reads was assessed with FastQC,^21^ and Trimmomatic^22^ was used for trimming adapter sequences and low-quality bases. Assemblies were performed using the A5 pipeline with default parameters.^23^ Sequence data are available under BioProject PRJNA603790.

### Identification of single nucleotide variants

<u>SNV calling was performed as in Han et al.^24^ The variant calling pipeline can be found at</u> https://github.com/Snitkin-Lab-Umich/variant_calling_pipeline. <u>To summarize, variant calling was performed with samtools^25^ using the reference genomes listed in Supplementary table 1</u>.

**Table 1.**
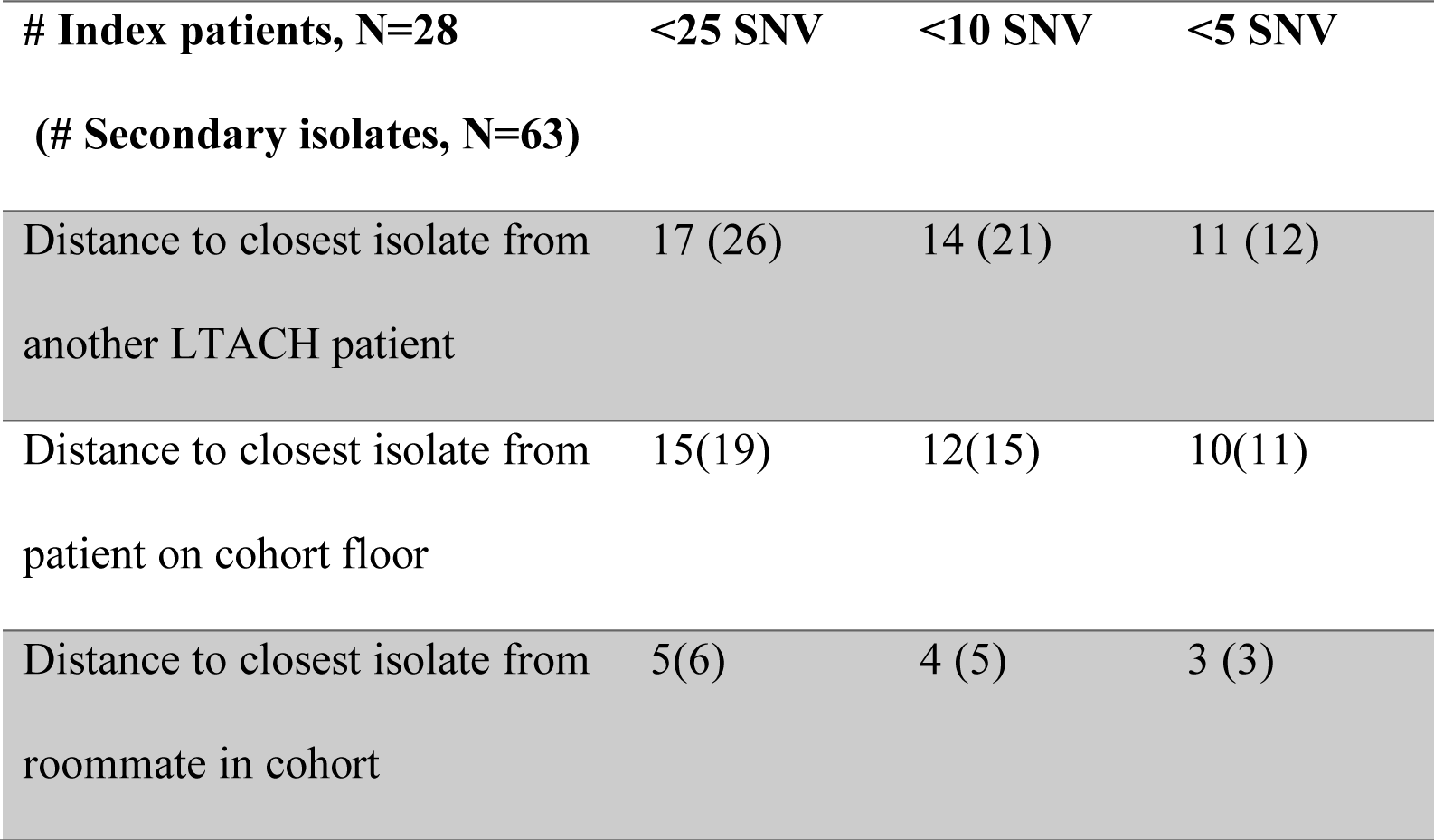
Frequency of strong genetic relationships between secondary isolates and isolates from other patients among patients whose primary isolate is most closely related to another patient’s isolate.

### Assessment of epidemiologically supported secondary acquisitions linked to other LTACH patients and roommates

Epidemiologically plausible donor patient isolates were defined as isolates collected before the recipient patient’s secondary isolate collection date. To account for acquisition potentially occurring between surveillance sampling dates, the positive donor time-frame for all analyses was defined starting on the date of the donor’s last negative swab before the collection date of the putative donor isolate.

The patient bed trace indicating the rooms patients were housed in during their LTACH stays was assessed to identify spatiotemporal exposures in shared patient rooms that plausibly facilitated secondary acquisition between roommates. Plausible secondary acquisitions linked to roommate exposures were defined as acquisitions between a donor and recipient patient who occupied the same room when the donor was considered positive for the putative donor isolate and prior to the collection date of the recipient’s secondary isolate.

### Genetic relationships between KPC-Kp isolates based on SNV distance

Pairwise distances were calculated from core and accessory genome single-nucleotide variants (SNVs) in whole-genome sequence alignments for each MLST represented by study isolates (Supplementary table 1). SNV distances were compared (1) between the first (primary) and later collected (secondary) isolates from the same index patient and (2) between secondary isolates from index patients and isolates from other plausible donor patients in the LTACH.

### Detection of resistance genes in whole genome sequences

Kleborate (https://github.com/katholt/Kleborate) was used to screen whole-genome sequence assemblies for presence of genes and mutations known to confer antibiotic resistance in *K pneumoniae*. We used a custom R script to expand antibiotic resistance gene alleles reported from Kleborate into gene presence absence profiles (Supplementary table 1), counting only the Kleborate-reported precise matching gene hits as being present or absent.

## Results

### Almost half of cohorted patients acquired secondary isolates of a new sequence type

We considered 127 ‘index’ patients, who were either positive at the start of the study or on first admission to the LTACH, for potential acquisition of secondary KPC-Kp strains during their stay. Although the original sampling strategy was not designed to track longitudinal colonization of KPC-Kp,^11^ there were 28 index patients who in addition to their 38 ‘primary’ isolates (earliest isolate) collected on admission or study start, also had 63 ‘secondary’ isolates collected later during their LTACH stays (**Figure 1**). Of the 101 isolates available from these index patients, we extracted quality WGS data from 99 isolates including 38 primary and 61 secondary isolates. While the majority of primary and secondary isolates were from the epidemic ST258 strain (55% of primary isolates, 57% of secondary isolates), a diversity of other multi-locus sequence types (MLSTs) was observed among both primary and secondary isolates (Supplemental Table 1). Secondary isolates were collected from patients a median of 89 days (range 1-310 days) after primary isolates. Evaluation of MLSTs of the primary and secondary KPC-Kp isolates provided support for secondary acquisition among cohorted patients, with 13 (46%) patients having a distinct secondary MLST that was not detected at admission.

**Figure 1.**
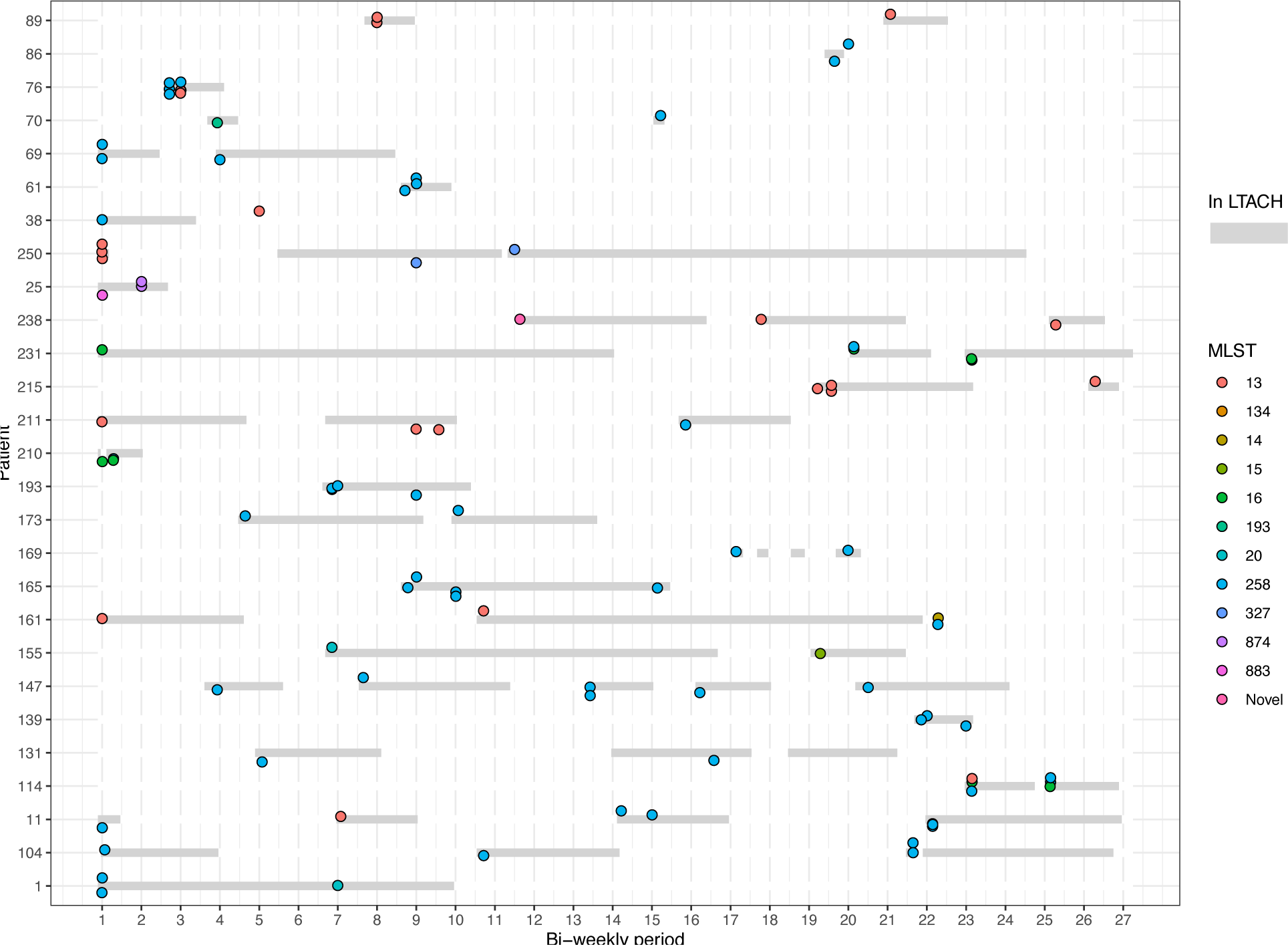
KPC-Kp isolates from convenience sample of patients who were positive at the study start or admission to the LTACH. Patients (N=28) have primary and secondary isolates that are from the same MLST, different MLST or both same and different MLST. Y-axis indicates patients, X-axis indicates bi-weekly time-periods during the study, circles indicate positive culture dates and are colored by the MLST of the isolate collected. Grey bars indicate when patients were in the LTACH.

### Genomic evidence of potential secondary acquisitions from other LTACH patients among admission-positive index patients

To assess genomic evidence of cross-transmission in the cohort we evaluated the fraction of patients whose secondary isolates were more closely related to another patient’s isolate than to their own primary isolate (**Figure 2**). Of the 28 index patients with one or more secondary isolates, 19 had a secondary isolate that was more closely related to another patient’s isolate than to their own primary isolate. Of those 19 patients, 17 had secondary isolates that were more closely related to an isolate from a patient with whom they overlapped on the cohort floor and 8 had secondary isolates that were more closely related to an isolate from a roommate. Plausible transmission in the cohort was further supported by extremely small SNV distances in most of these cases, with 12 patients’ isolates being within 10 SNVs of another patient’s isolate on the cohort floor and 4 patients’ isolates being within 10 SNVs of an isolate from a roommate (**Table 1**).

**Figure 2.**
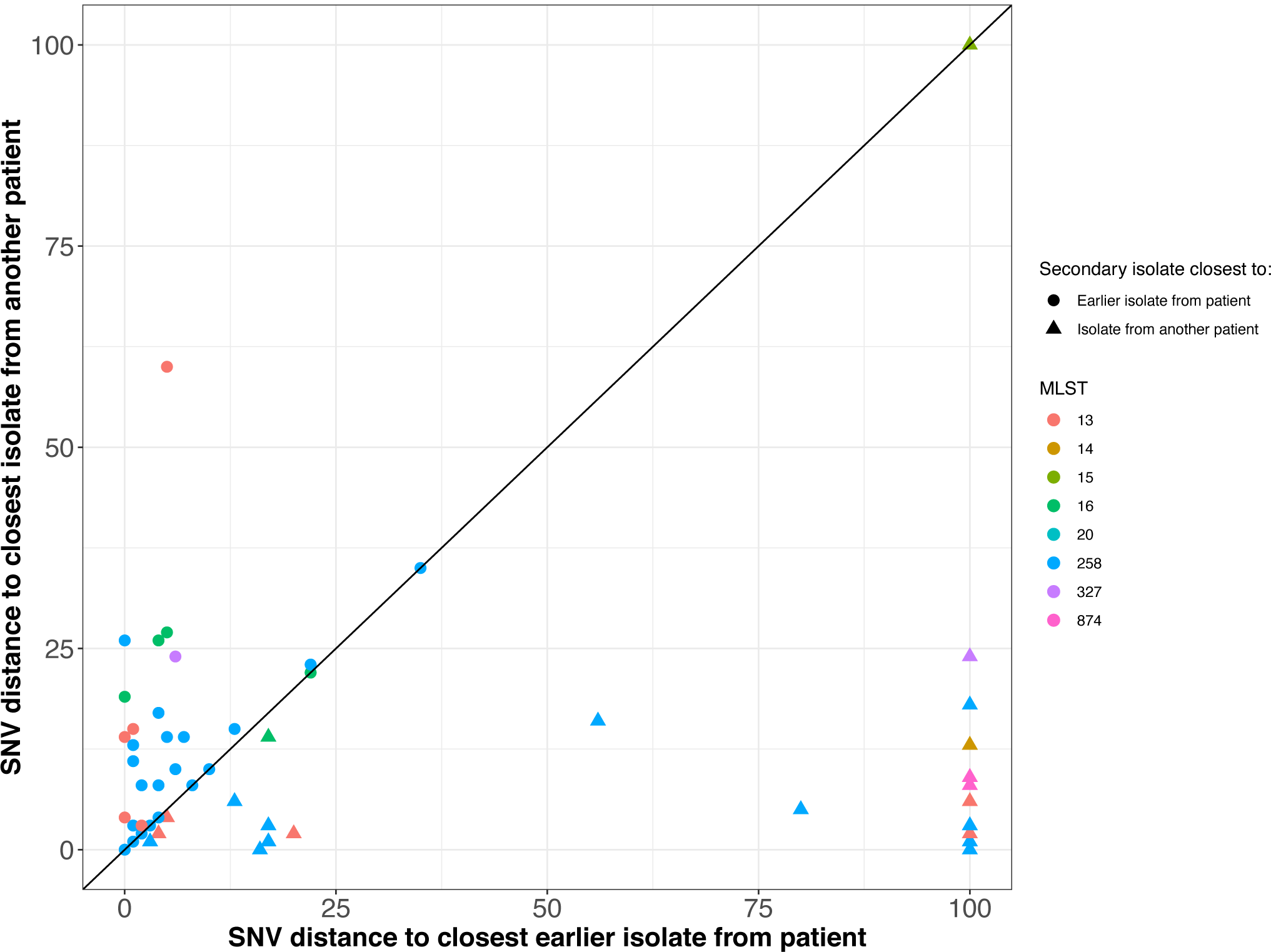
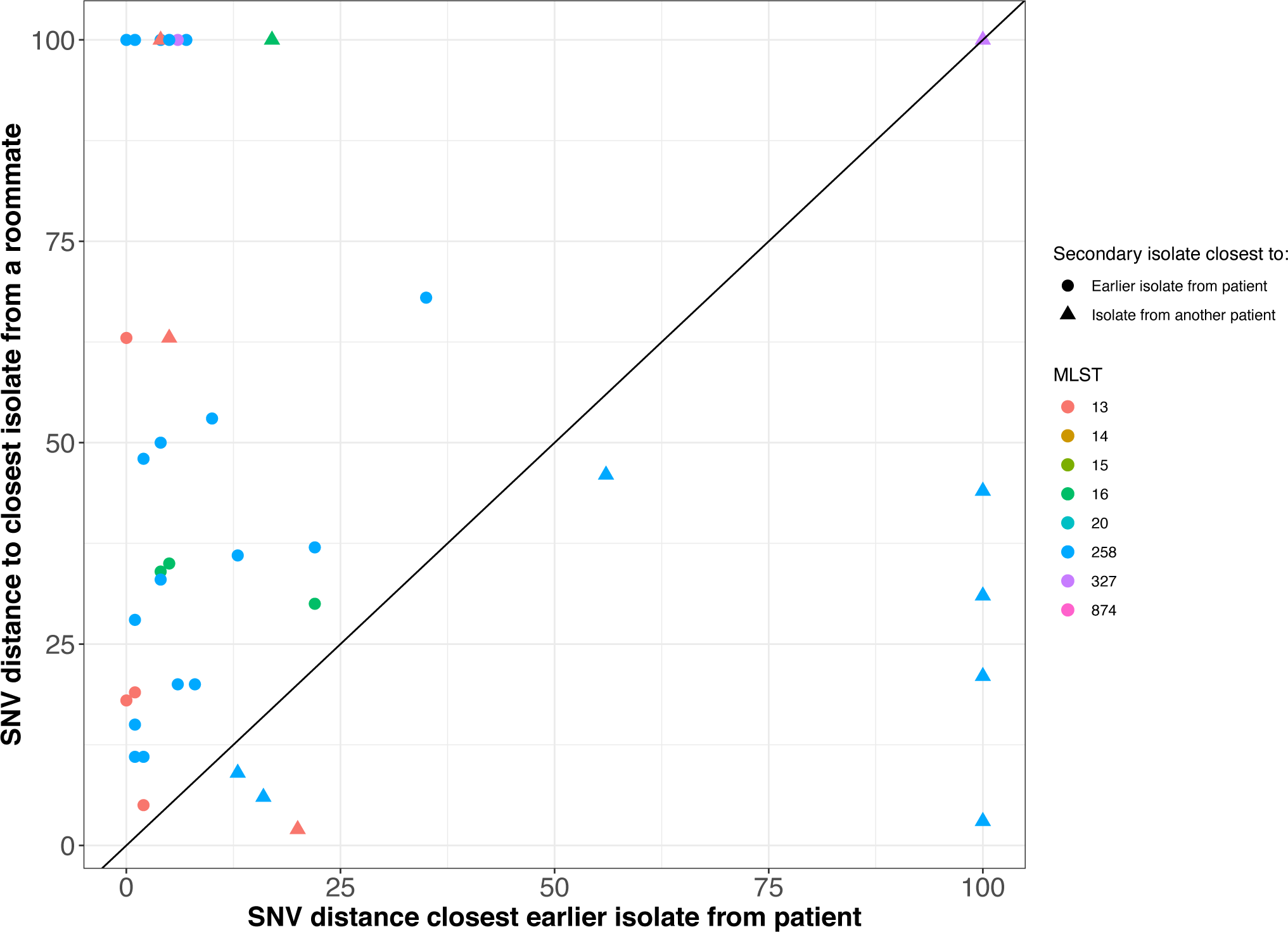
Genetic relationship between a patient’s primary and secondary isolates compared to isolates from other patients in the LTACH and room cohorts. Pairwise SNV distance between secondary isolates and closest primary isolate from the same patient compared to closest related isolate from **A**. another patient in the facility or **B**. a cohorted roommate. Diagonal line separates secondary isolates that are more closely related to primary isolates from the same patient (above the diagonal) or to another patient’s isolate (below the diagonal). Colors indicate the MLST of the secondary isolate. Circles indicate the closest genetic relative to the isolate by SNV distance is from the same patient (e.g. the patient’s own primary isolate) while triangles indicate that the closest relative was isolated from another patient. Comparison of isolates from different MLSTs or >100 SNVs are collapsed into the >100 SNV category for plot visualization purposes.

### Patients accumulate diverse antibiotic resistance genes in association with acquisition of a secondary KPC-Kp isolate

There is an abundance of molecular and genomic evidence that members of the same bacterial species, including KPC-Kp, can vary extensively in the arsenal of antibiotic resistance genes encoded in their chromosomes and plasmids.^12,26,27^ To determine whether secondary acquisitions resulted in increased antibiotic resistance potential we examined whether patients with high-confidence putative transmission links (<10 SNVs to another patient’s isolate and >10 SNVs from their own primary isolate) acquired additional unique resistance genes in their secondary isolate. As compared to a patient’s primary isolate, secondary isolates contributed a median of 2.5 additional antibiotic resistance genes beyond the primary isolate (minimum 0, maximum 10 additional resistance genes) (**Table 2**). In total, additional resistance genes were gained in 12 of the 14 patients whose secondary isolates had strong genomic links to isolates from other patients, including 3 patients whose secondary isolates were linked to patients with whom they had shared a cohort room prior to secondary isolate acquisition (**Figure 3**, supplementary table 1,Patients with unlinked secondary isolates accumulated fewer additional resistance genes (median 0, minimum 0, maximum 2 additional resistance genes) (supplementary figure 1).). This finding supports the hypothesis that these closely related isolates (<10 SNVs) represented primary isolates that accrued mutations over the course of prolonged colonization rather than that patients acquired a secondary KPC-Kp strain via transmission from another patient.

**Table 2.**
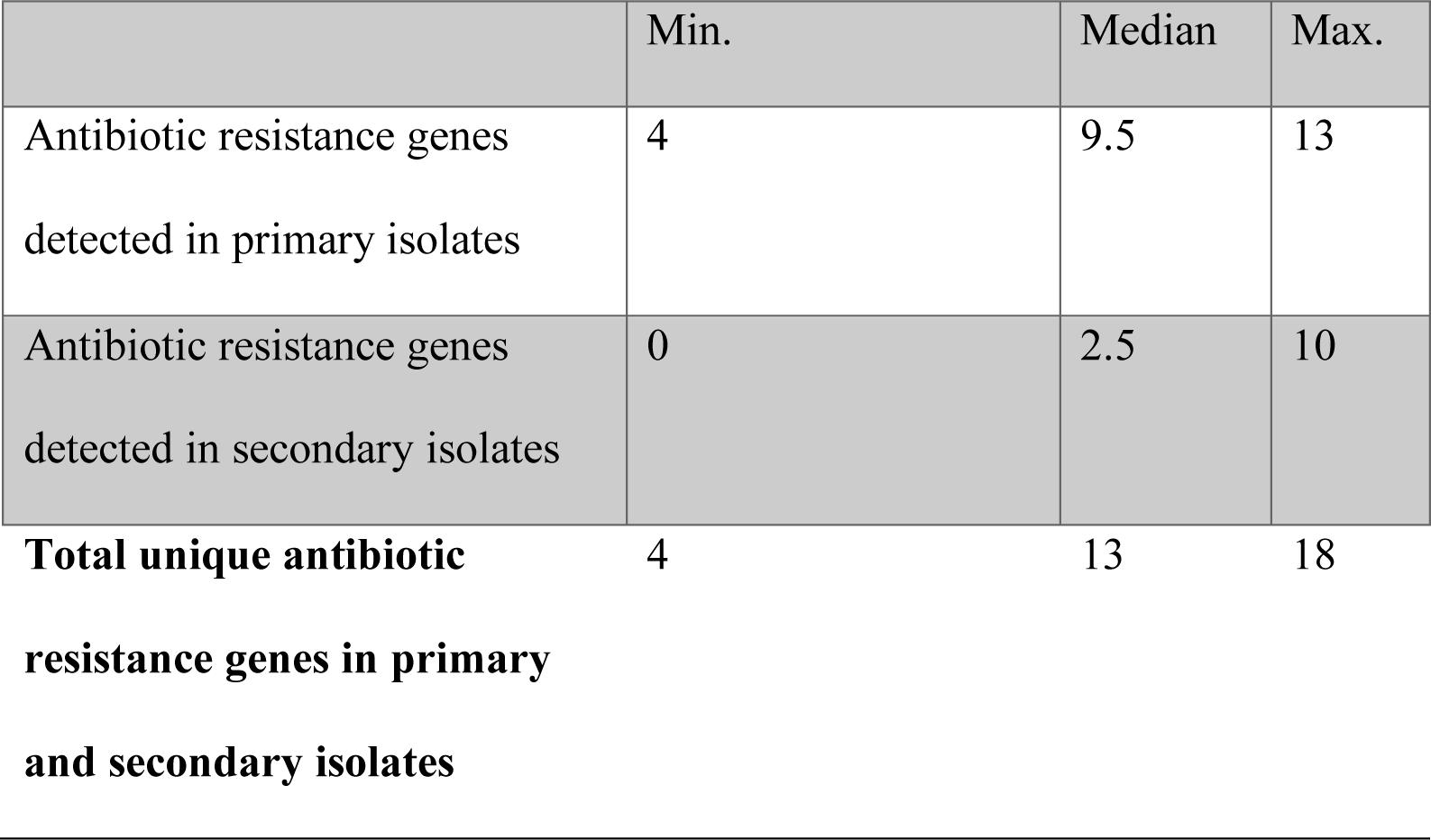
Summary of antibiotic resistance genes among primary, secondary and all isolates from index patients whose secondary isolate is most closely related to another patient’s isolate.

**Figure 3.**
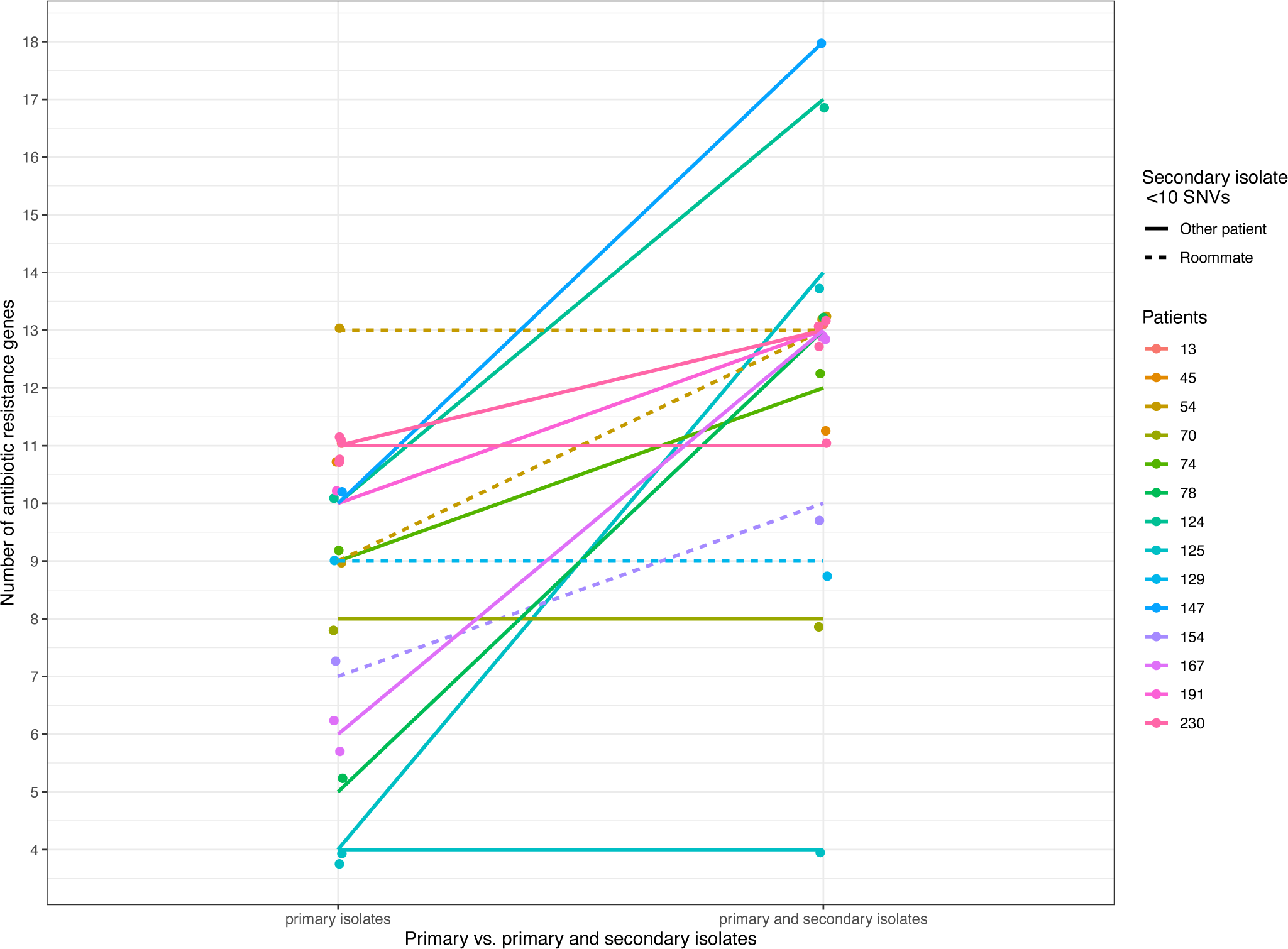
Number of antibiotic resistance genes detected in genomes from primary isolates compared to primary and secondary isolates from index patients whose secondary isolates are linked with high confidence (<10 SNVs) to isolates from other patients in the LTACH. Y-axis indicates number of unique resistance genes detected with Kleborate (See methods, Supplementary table 1), X-axis indicates number of unique antibiotic resistance genes detected among primary (left) and primary and secondary isolates (right). Colors distinguish patients. Dashed lines indicate patients whose secondary isolate is within 10 SNVs of an isolate from a cohorted roommate.

## Discussion

Cohorting patients who are colonized or infected with MDROs is an effective strategy to reduce the risk of MDRO transmission to uncolonized patients. However, little attention has been paid to the potential for cohorted patients themselves to acquire secondary resistant strains through exposure to the high colonization pressure of MDROs within cohorts. Secondary strain acquisition may be particularly important in endemic settings where the MDRO for which patients are cohorted, e.g. CRE, may comprise a heterogeneous group of bacteria with varying genetic potential. In order to investigate this risk, we performed a genomic epidemiologic investigation of a longitudinal, convenience sample of KPC-Kp isolates from patients on cohort floors in a LTACH. We found strong evidence of cross-transmission within cohorts, with secondary acquired isolates often harboring antibiotic resistance genes not found within a patient’s primary isolate.

Our finding that secondary isolates carry antibiotic resistance potential that is distinct from that found in patients’ primary isolates is noteworthy because it suggests that multiple strain acquisition could increase risk of treatment failure. Acquisition of a secondary strain that is resistant to antibiotics to which the primary strain was susceptible could be particularly problematic for highly resistant organisms like KPC-Kp, which already have limited treatment options. For example, colistin/polymyxin E is a last-resort drug that is used to treat severe multidrug-resistant gram negative infections, such as those due to KPC-Kp.^28–31^ In our study, one patient plausibly acquired a secondary isolate with predicted colistin resistance that was linked within 25 SNVs of another LTACH patient’s isolate (supplementary table 1). As colonization is a major risk factor for KPC-Kp infection,^32–34^ and infections are thought to arise primarily from the patient’s colonizing strain, ^35^ the acquisition of a colistin resistant isolate could limit efficacious treatment options and in turn increase mortality risk. ^31,36^ In addition to the potential risks to multiply colonized patients, the acquisition of strains with different resistance arsenals provides an opportunity for horizontal gene exchange and the accumulation of resistance within a single transmissible strain.^19,20,37^ Moreover, harboring genetically diverse strains creates an opportunity for resistance alleles to find their way to strains with other clinically relevant characteristics, such as hyper-virulence^13,38–40^ or epidemic potential.^39^ Additional risk to patients could stem from the fact that different strains of the same pathogen often carry different virulence genes.^37^ Virulence factor differences in acquired strains may predispose patients to developing infections of different types and severity.^37,38^

In addition to potentially making infections more difficult to treat, acquisition of secondary strains could also increase a patient’s time at risk of infection by prolonging the total period of colonization. All of these potential adverse consequences of multiple strain colonization emphasize the importance of protecting previously colonized patients from secondary acquisition and for healthcare providers to adhere to infection prevention protocols, even when caring for patients in cohort locations.

Our study has several limitations. First, we studied a convenience sample which inherently precludes systematic calculation of risk. Second, we conducted limited sequencing of multiple clones from the same sample—a single representative of unique morphologies observed in each sample, primarily a single clone per sample--thus hindering our ability to know if a patient was simultaneously colonized with multiple strains (e.g. colonized by both their primary and secondary strains at the same time). These sampling limitations also prevent us from determining if patients remain colonized with their primary strain when they become colonized with their secondary strain, or if colonization with both strains persists. Thus, it is possible that cohort patients entered the facility already colonized with multiple strains, and that patients did not acquire their secondary strains in the cohort. While we cannot definitively rule out this possibility, the acquisition of secondary strains in the LTACH is supported by the finding that 14 of the 28 patients with secondary isolates had strong genomic links (< 10 SNVs) to other LTACH patients. In total, these 14 strong genomic linkages account for 50% of the 28 index patients with multiple isolates available and 11% of the 127 index patients in the full study.

In summary, our study provides strong evidence for cross-transmission of KPC-Kp strains within a KPC-Kp-positive cohort, with accumulation of new antibiotic resistance genes by patients who acquire secondary KPC-Kp strains. Whether acquisition of multiple KPC-Kp strains increases risk of adverse patient outcomes needs to be studied further. In the meantime, we recommend robust adherence to infection prevention precautions within KPC-Kp cohorts to reduce the risk of within-cohort cross-transmission of KPC-Kp strains.

## Data Availability

Sequence data are available under BioProject PRJNA603790.

## Acknowledgements

We thank the patients and staff of the Long-term acute-care hospital (LTACH) for their gracious participation in this study; Ali Pirani for bioinformatics support and members of the Snitkin lab and the Rush University/University of Michigan genomics working group for critical review of the manuscript. All authors (S.E.H, M.K.H, K.L, R.D.Y, R.A.W, M.Y.L, and E.S.S) report no conflicts of interest.

## Financial support

This work was supported by CDC U54 CK00016 04S2 and CDC U54 CK000481. S.E.H was supported by the University of Michigan NIH Training Program in Translational Research T32-GM113900 and the University of Michigan Rackham pre-doctoral fellowship.

## Notes

### Competing Interest Statement

The authors have declared no competing interest.

